# Factors Associated with Hospitalization and Disease Severity in a Racially and Ethnically Diverse Population of COVID-19 Patients

**DOI:** 10.1101/2020.06.25.20137323

**Authors:** Angelico Mendy, Senu Apewokin, Anjanette A. Wells, Ardythe L. Morrow

## Abstract

**Background:** The coronavirus disease (COVID-19) first identified in Wuhan in December 2019 became a pandemic within a few months of its discovery. The impact of COVID-19 is due to both its rapid spread and its severity, but the determinants of severity have not been fully delineated.

**Objective:** Identify factors associated with hospitalization and disease severity in a racially and ethnically diverse cohort of COVID-19 patients.

**Methods:** We analyzed data from COVID-19 patients diagnosed at the University of Cincinnati health system from March 13, 2020 to May 31, 2020. Severe COVID-19 was defined as admission to intensive care unit or death. Logistic regression modeling adjusted for covariates was used to identify the factors associated with hospitalization and severe COVID-19.

**Results:** Among the 689 COVID-19 patients included in our study, 29.2% were non-Hispanic White, 25.5% were non-Hispanic Black, 32.5% were Hispanic, and 12.8% were of ‘Other’ race/ethnicity. About 31.3% of patients were hospitalized and 13.2% had severe disease. In adjusted analyses, the sociodemographic factors associated with hospitalization and/or disease severity included older age, non-Hispanic Black or Hispanic race/ethnicity (compared to non-Hispanic White), and smoking. The following comorbidities: diabetes, hypercholesterolemia, asthma, COPD, chronic kidney disease, cardiovascular diseases, osteoarthritis, and vitamin D deficiency were associated with hospitalization and/or disease severity. Hematological disorders such as anemia, coagulation disorders, and thrombocytopenia were associated with both hospitalization and disease severity.

**Conclusion:** This study confirms race and ethnicity as predictors of severe COVID-19. It also finds clinical risk factors for hospitalization and severe COVID-19 not previously identified such a vitamin D deficiency, hypercholesterolemia, osteoarthritis, and anemia.

## INTRODUCTION

In December 2019, an outbreak of severe acute respiratory syndrome coronavirus-2 (SARS-CoV-2) pneumonia appeared in Wuhan and rapidly spread throughout the world causing more than 8 million infections and close to 450,000 deaths by June 2020.^1,2^ Similar to previous coronaviruses, the newly identified virus is highly contagious; it is transmitted primarily through droplets and causes major outbreaks in the absence of adequate control measures.^3^ The manifestations of the coronavirus disease 2019 (COVID-19) caused by SARS-CoV-2 are widely variable. The infection is asymptomatic in some individuals, while in others, it causes symptoms ranging from dry cough and dyspnea to severe pneumonia with respiratory failure requiring admission in intensive care unit (ICU) and leading to death in severe cases.^4,5^

To date, a number of studies have investigated the predictors of severe COVID-19. Nevertheless, the sociodemographic and clinical factors that influence this disease course have not been fully defined.^6-12^ Whether asthma, a common respiratory illness affecting 1 in 12 American adults, is associated with disease severity in COVID-19 is still a matter of debate.^13-17^ Other important clinical factors remain understudied. For example, vitamin D deficiency affects 1 in 4 American adults, has an important immunologic role and has been suggested to increase the risk of SARS-CoV-2 infection.^18^ However, vitamin D deficiency has not been analyzed as a risk factor for severe COVID-19 among patients infected with SARS-CoV-2. Another understudied clinical factor is hypercholesterolemia, which affects 1 in 8 American adults; whether this condition has been reported to be associated with severe COVID-19 has not been analyzed in the published literature.^19,20^ Moreover, most of the research on the potential factors for severe COVID-19 have been conducted in China among Chinese patients which may limit their generalizability to multi-ethnic populations.^6-8,10^ The reports that have included a racially diverse study sample are scant^9,11,12,21^ To address this gap in the literature, we aimed to determine the factors associated with hospitalization and disease severity in a racially and ethnically diverse population of COVID-19 patients by including some comorbidities missing in previous studies.

## METHODS

### Data Source

Data were extracted from the electronic medical record system for all COVID-19 patients diagnosed at the University of Cincinnati health system (UC Health) between March 13, 2020 to May 31, 2020. UC Health consists of 4 hospitals located in the Cincinnati metropolitan area and primary care and specialty clinics located in the states of Ohio, Kentucky, and Indiana. COVID-19 diagnosis was defined as a positive nasopharyngeal reverse transcriptase polymerase chain reaction test for SARS-CoV-2. A total of 691 patients were diagnosed with COVID-19 and after exclusion of two patients who did not report their sex, 689 patients were included in our study.

### Variables

Age at the time of COVID-19 diagnosis was calculated using patients date of birth. Sex, race/ethnicity, and smoking were self-reported. Comorbidities, as well as hematological disorders were defined using the 10^th^ revision of the International Classification of Diseases (ICD10) codes. The following comorbidities were well characterized in our data and were included in the study: obesity (E66), diabetes (E10 and E11), pure hypercholesterolemia (E78.0), asthma (J45), chronic obstructive pulmonary disease (COPD) (J44), chronic kidney disease (N18), cardiovascular disease (I00-I99), neoplasm or a history of neoplasm (C00-D49), osteoarthritis (M15-M19), and vitamin D deficiency (E55). The hematological disorders we included were anemia (D50-D53 for nutritional anemia and D55-D59 for hemolytic anemia), coagulation defects, purpura and other hemorrhagic conditions (D65-D69), and thrombocytopenia (D69.6).

### Definition of Outcome Variables

Hospitalization was defined as admission to the hospital for at least 24 consecutive hours. Disease severity was defined as admission to ICU and/or death during hospitalization. We also estimated the length of hospital stay among the COVID-19 patients who were hospitalized and successfully discharged. Those who died during hospitalization were excluded from the length of hospital stay analysis.

### Statistical Analysis

Descriptive analyses were performed to summarize the characteristics of the COVID-19 patients overall and by hospitalization or disease severity status. Chi-square or Fisher’s exact tests were used to estimate P-values for differences. The differences in the length of hospital stay were evaluated by means of Wilcoxon Rank Sum tests. The covariates-adjusted odds ratios (OR) and 95% confidence intervals (CI) for the associations of the patients’ characteristics and comorbidities with hospitalization and disease severity were estimated by means of logistic regression analyses. We also performed multinomial logistic regressions to further stratify disease severity into admission to ICU and death. To identify the factors associated with length of hospital stay, we fitted generalized linear models with a gamma distribution and a log link function which is similar in shape to the log-normal distribution and is robust to the outcome of length of hospital stay that was right skewed.^22^ The diagnostic plots for the generalized linear models showed that the assumptions were reasonably met and the regression coefficients (β) along with the 95% CI for the associations were reported. All the models were adjusted for age, sex, race/ethnicity, and cigarette smoking. The analyses were performed in SAS Version 9.4 (SAS Institute, Cary, NC) and two-sided p-values <0.05 were considered statistically significant in all analyses.

## RESULTS

### Description of Study Population

The 689 COVID-19 patients included in our study had a median age of 49.5 years (IQR: 35.2, 67.5) and 53.0% were male. The race/ethnicity of study patients was 29.2% non-Hispanic White, 25.5% non-Hispanic Black, 32.5% Hispanic, and 12.8% patients of ‘Other’ race/ethnicity. Cardiovascular disease was the most common comorbidity (49.5%). The other comorbidities included diabetes (24.7%), neoplasm or history of neoplasm (19.7%), obesity (18.6%), osteoarthritis (14.2%), vitamin D deficiency (12.9%), chronic kidney disease (11.8%), asthma (10.2%), COPD (8.8%), and pure hypercholesterolemia (2.9%). Hematological disorders were reported in 25.5% of patients for anemia, 8.0% for coagulation defect, and 5.4% thrombocytopenia (Table 1).

**Table 1:** Characteristics of study participants (N = 689)

| Characteristics | All<br>Participants | Hospitalization |  |  | Severe disease |  |  |
| --- | --- | --- | --- | --- | --- | --- | --- |
|  |  | No | Yes | P-value | No | Yes | P-value |
| Prevalence, N (%) | 689 (100.0) | 473 (68.7) | 216 (31.3) |  | 598 (86.8) | 91 (13.2) |  |
| **Socio-demographic characteristics** |  |  |  |  |  |  |  |
| Age, years, median (SE) | 49.5 (1.3) | **44.38 (1.2)** | **60.3 (2.0)** | **< 0.001** | **47.2 (1.3)** | **60.5 (3.4)** | **0.001** |
| Male sex, N (%) | 365 (53.0) | 245 (51.8) | 120 (55.6) | 0.359 | 319 (53.3) | 46 (50.5) | 0.619 |
| Race/ethnicity, N (%) |  |  |  | **< 0.001** |  |  | **< 0.001** |
| Non-Hispanic Whites | 201 (29.2) | **138 (29.2)** | **63 (29.2)** |  | **183 (30.6)** | **23 (25.3)** |  |
| Non-Hispanic Blacks | 176 (25.5) | **91 (19.2)** | **85 (39.3)** |  | **135 (22.6)** | **43 (47.3)** |  |
| Hispanics | 224 (32.5) | **169 (35.7)** | **55 (25.5)** |  | **187 (31.3)** | **23 (23.3)** |  |
| Other | 88 (12.8) | **75 (15.9)** | **13 (6.0)** |  | **93 (15.6)** | **2 (2.2)** |  |
| Past and current smoker,<sup>a</sup> N (%) | 170 (24.7) | **84 (17.8)** | **86 (39.8)** | **< 0.001** | **133 (22.2)** | **37 (40.7)** | **< 0.001** |
| **Comorbidities** |  |  |  |  |  |  |  |
| Obesity, N (%) | 128 (18.6) | **75 (15.9)** | **53 (24.5)** | **0.007** | **104 (17.4)** | **24 (26.4)** | **0.040** |
| Diabetes, N (%) | 170 (24.7) | **76 (16.1)** | **94 (43.5)** | **< 0.001** | **124 (20.7)** | **46 (50.5)** | **< 0.001** |
| Pure hypercholesterolemia, N (%) | 20 (2.9) | **2 (0.4)** | **18 (8.3)** | **< 0.001** | **10 (1.7)** | **10 (11.9)** | **< 0.001** |
| Asthma, N (%) | 70 (10.2) | **34 (7.2)** | **36 (16.7)** | **< 0.001** | **47 (7.9)** | **23 (25.3)** | **< 0.001** |
| COPD, N (%) | 61 (8.8) | **23 (4.9)** | **38 (17.6)** | **< 0.001** | **38 (6.3)** | **23 (27.4)** | **< 0.001** |
| Chronic kidney disease, N (%) | 81 (11.8) | **24 (5.1)** | **57 (26.4)** | **< 0.001** | **46 (7.7)** | **35 (38.5)** | **< 0.001** |
| Cardiovascular disease, N (%) | 341 (49.5) | **174 (36.8)** | **167 (77.3)** | **< 0.001** | **265 (44.3)** | **76 (83.5)** | **< 0.001** |
| Neoplasm or history of neoplasm, N (%) | 136 (19.7) | **71 (15.0)** | **65 (30.1)** | **< 0.001** | **106 (17.7)** | **30 (33.0)** | **< 0.001** |
| Osteoarthritis, N (%) | 105 (14.2) | **46 (9.7)** | **59 (27.3)** | **< 0.001** | **78 (13.0)** | **27 (29.7)** | **< 0.001** |
| Vitamin D Deficiency, N (%) | 89 (12.9) | **44 (9.3)** | **45 (20.8)** | **< 0.001** | **66 (11.0)** | **23 (25.3)** | **< 0.001** |
| **Hematological disorders** |  |  |  |  |  |  |  |
| Anemia, N (%) | 176 (25.5) | **82 (17.3)** | **94 (43.5)** | **< 0.001** | **123 (20.6)** | **53 (58.2)** | **< 0.001** |
| Coagulation defect, N (%) | 55 (8.0) | **15 (3.2)** | **40 (18.5)** | **< 0.001** | **29 (4.8)** | **26 (28.6)** | **< 0.001** |
| Thrombocytopenia, N (%) | 37 (5.4) | **9 (1.9)** | **28 (13.0)** | **< 0.001** | **18 (3.0)** | **19 (20.9)** | **< 0.001** |

Abbreviations: SE, standard error; COPD, chronic obstructive pulmonary disease.
P-values for difference calculated using chi-square or Fisher's exact tests.
Severe disease defined as admission to intensive care unit or death.
<sup>a</sup> 22.8% of participants had missing data on smoking

Two-hundred-sixteen (31.3%) of COVID-19 patients were hospitalized and 91 (13.2%) had the severe form of the disease. The COVID-19 patients who were hospitalized and/or had severe disease tended to be older, to be non-Hispanic Black, and to be past or current smokers, compared to those who were not hospitalized or free of severe disease. Hospitalized patients and those with severe disease also tended to have comorbidities such as obesity, diabetes, pure hypercholesterolemia, asthma, COPD, chronic kidney disease, cardiovascular disease, neoplasm or history of neoplasm, osteoarthritis, vitamin D deficiency, and hematological disorders (anemia, coagulation defect, and thrombocytopenia) (Table 1).

Among the COVID-19 patients who were hospitalized and survived (N=191), the median length of hospital stay was 6.91 days (IQR: 3.27, 11.56) (Table 2). This duration was longer in adults aged 60 years or older compared with younger individuals, in non-Hispanic Whites and non-Hispanic Blacks, in patients with diabetes, asthma, COPD, chronic kidney disease, cardiovascular disease, neoplasm or history of neoplasm, vitamin D deficiency, or hematological disorders (anemia, coagulation defect, and thrombocythemia) (Table 2).

**Table 2:** Length of hospital stay (in days) overall and by characteristics of hospitalized study patients (N = 191)

| Characteristics | N | Median (IQR) | P-value |
| --- | --- | --- | --- |
| All participants | 191 | 6.91 (3.27, 11.56) |  |
| **Sociodemographic characteristics** |  |  |  |
| Age groups |  |  | **0.005** |
| <60 years | **92** | **5.25 (2.50, 11.08)** |  |
| ≥60 years | **99** | **7.76 (5.01, 12.44)** |  |
| Sex |  |  | 0.056 |
| Female | 84 | 5.77 (3.01, 9.76) |  |
| Male | 107 | 7.39 (4.23, 14.45) |  |
| Race/ethnicity |  |  | **0.001** |
| Non-Hispanic Whites | **54** | **8.25 (4.58, 15.47)** |  |
| Non-Hispanic Blacks | **72** | **7.76 (4.36, 11.93)** |  |
| Hispanics | **54** | **4.62 (2.35, 8.68)** |  |
| Other | **11** | **4.95 (2.68, 5.99)** |  |
| Smoking<sup>a</sup> |  |  | 0.117 |
| Never smoker | 88 | 6.08 (3.20, 10.89) |  |
| Past and current smoker | 76 | 7.93 (4.00, 14.35) |  |
| **Comorbidities** |  |  |  |
| Obesity |  |  | 0.799 |
| No | 143 | 6.70 (3.33, 12.00) |  |
| Yes | 48 | 6.93 (3.25, 10.72) |  |
| Diabetes |  |  | **< 0.001** |
| No | **109** | **5.37 (2.94, 8.79)** |  |
| Yes | **82** | **8.73 (4.51, 15.10)** |  |
| Pure hypercholesterolemia |  |  | 0.065 |
| No | 176 | 6.66 (3.22, 11.21) |  |
| Yes | 15 | 9.61 (5.30, 14.97) |  |
| Asthma |  |  | **< 0.001** |
| No | 158 | **5.77 (3.12, 10.08)** |  |
| Yes | 33 | **11.05 (7.25, 16.010)** |  |
| COPD |  |  | **0.002** |
| No | 158 | **6.13 (3.12, 10.47)** |  |
| Yes | 33 | **10.36 (5.23, 16.84)** |  |
| Chronic kidney disease |  |  | **0.012** |
| No | 146 | **5.84 (3.03, 10.47)** |  |
| Yes | 45 | **8.57 (5.11, 15.17)** |  |
| Cardiovascular disease |  |  | **< 0.001** |
| No | 142 | **4.76 (2.50, 8.01)** |  |
| Yes | 49 | **7.76 (4.22, 14.55)** |  |
| Neoplasm or history of neoplasm |  |  | **0.004** |
| No | 135 | **5.81 (3.00, 10.290)** |  |
| Yes | 56 | **8.28 (5.12, 16.00)** |  |
| Osteoarthritis |  |  | 0.137 |
| No | 139 | 6.19 (3.04, 11.03) |  |
| Yes | 52 | 7.17 (4.96, 14.35) |  |
| Vitamin D Deficiency |  |  | **< 0.001** |
| No | 151 | **5.79 (3.01, 10.04)** |  |
| Yes | 40 | **10.23 (5.77, 18.12)** |  |
| **Hematological disorders** |  |  |  |
| Anemia |  |  | **0.003** |
| No | 110 | **5.66 (2.98, 8.82)** |  |
| Yes | 81 | **8.32 (4.97, 15.82)** |  |
| Coagulation defect |  |  | **0.012** |
| No | 160 | **6.13 (3.24, 10.72)** |  |
| Yes | 31 | **10.56 (5.05, 21.75)** |  |
| Thrombocytopenia |  |  | **0.003** |
| No | 171 | **6.21 (3.22, 10.84)** |  |
| Yes | 20 | **11.04 (5.77, 28.21)** |  |

Abbreviations: IQR, interquartile range COPD, chronic obstructive pulmonary disease; N, number of participants in subgroups. P-values for difference calculated using Wilcoxon rank tests.

### Factors Associated with Hospitalization

In adjusted analysis, the sociodemographic characteristics associated with hospitalization were age (OR: 1.36, 95% CI: 1.22, 1.51 per 10-year increase), being non-Hispanic Black (OR: 2.23, 95% CI: 1.41, 3.53) or Hispanic (OR: 1.91, 95% CI: 1.11, 3.29) (compared to being non-Hispanic White), and smoking (OR: 2.01, 95% CI: 1.32, 3.06). The comorbidities associated with higher odds of hospitalization included diabetes (OR: 2.62, 95% CI: 1.75, 3.90), pure hypercholesterolemia (OR: 9.30, 95% CI: 2.02, 42.74), asthma (OR: 1.92, 95% CI: 1.10, 3.35), chronic kidney disease (OR: 3.47, 95% CI: 1.99, 6.07), cardiovascular disease (OR: 4.39, 95% CI: 2.75, 7.01), osteoarthritis (OR: 1.95, 95% CI: 1.19, 3.19), and vitamin D deficiency (OR: 1.77, 95% CI: 1.07, 2.93). The hematological disorders (anemia [OR: 2.59, 95% CI: 1.72, 3.91], coagulation defect [OR: 4.90, 95% CI: 2.52, 9.56], and thrombocytopenia [OR: 6.03, 95% CI: 2.64, 13.77]) were also associated with higher prevalence of hospitalization (Table 3).

**Table 3:**
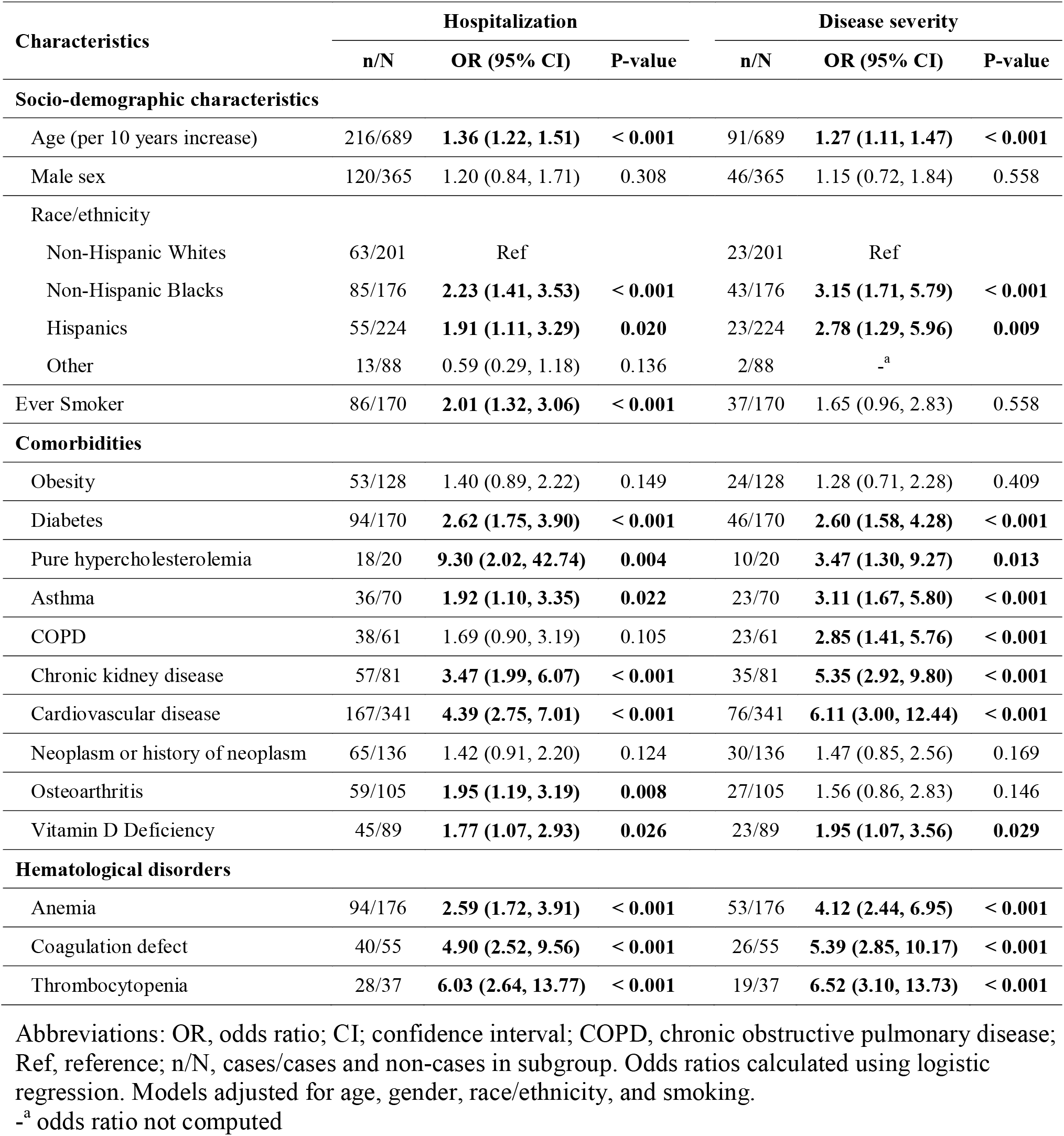
Risk Factors for hospitalization and disease severity in adjusted analysis (N = 689)

### Factors Associated with Disease Severity

#### Disease severity

The sociodemographic characteristics associated with severe COVID-19, after adjustment for covariates were age (OR: 1.27, 95% CI: 1.11, 1.47) and being non-Hispanic black (OR: 3.15, 95% CI: 1.71, 5.79) or Hispanic (OR: 2.78, 95% CI: 1.29, 5.96) compared to non-Hispanic White. Among the studied comorbidities, the odds of severe disease was higher in patients with diabetes (OR: 2.60, 95% CI: 1.58, 4.28), pure hypercholesterolemia (OR: 3.47, 95% CI: 1.30, 9.27), asthma (OR: 3.11, 95% CI: 1.67, 5.80), COPD (OR: 2.85, 95% CI: 1.41, 5.76), chronic kidney disease (OR: 5.35, 95% CI: 2.92, 9.80), cardiovascular disease (OR: 6.11, 95% CI: 3.00, 12.44), and vitamin D deficiency (OR: 1.95, 95% CI: 1.07, 3.56). As for hospitalization, hematological disorders (anemia [OR: 4.12, 95% CI: 2.44, 6.95], coagulation defect [OR: 5.39, 95% CI: 2.85, 10.17], and thrombocytopenia [OR: 6.52, 95% CI: 3.10, 13.73]) had a strong positive association with severe COVID-19 (Table 3).

#### Admission to ICU and Death

In multinomial analysis stratifying COVID-19 severity into admission to ICU and death, being non-Hispanic Black compared to being non-Hispanic White was the only sociodemographic characteristic associated with both admission to ICU (OR: 3.32, 95% CI: 1.56, 7.07) and death (OR: 3.44, 95% CI: 1.32, 9.00). Age was associated with increased death (OR: 1.94, 95% CI: 1.47, 2.58 per 10-year increase), while Hispanics (OR: 3.44, 95% CI: 1.42, 8.34) and smokers (OR: 2.34, 95% CI: 1.23, 4.46) had higher odds of admission to ICU.

Among comorbidities, odds of admission to ICU and death were both higher in patients with chronic kidney disease (OR: 5.63, 95% CI: 2.72, 11.64 for admission to ICU and OR: 4.48, 95% CI: 1.81, 11.08). The odds of admission to ICU were higher in diabetes (OR: 3.31, 95% CI: 1.84, 5.96), pure hypercholesterolemia (OR: 3.77, 95% CI: 1.25, 11.36), asthma (OR: 4.33, 95% CI: 2.18, 8.58), COPD (OR: 4.26, 95% CI: 1.87, 9.77), cardiovascular disease (OR: 5.59, 95% CI: 2.57, 12.14), and vitamin D deficiency (OR: 2.55, 95% CI: 1.28, 5.08).

The studied hematological disorders were associated with both admission to ICU (OR: 5.19, 95% CI: 2.80, 9.65 for anemia, OR: 4.67, 95% CI: 2.28, 9.57 for coagulation defects, and OR: 5.14, 95% CI: 2.18, 12.09 for thrombocytopenia) and death (OR: 2.58, 95% CI: 1.05, 6.38 for anemia, OR: 8.81, 95% CI: 3.11, 24.98 for coagulation defect, and OR: 14.12, 95% CI: 4.54, 43.84 for thrombocytopenia) (Table 4).

**Table 4:** Factors associated with admission to ICU and death (N = 689)

| Characteristics | Admission to ICU |  |  | Death |  |  |
| --- | --- | --- | --- | --- | --- | --- |
|  | n/N | OR (95% CI) | P-value | n/N | OR (95% CI) | P-value |
| **Sociodemographic characteristics** |  |  |  |  |  |  |
| Age, per 10 year-increase | 65/689 | 1.08 (0.92, 1.28) | 0.340 | 26/689 | **1.94 (1.47, 2.58)** | **< 0.001** |
| Male sex | 32/365 | 0.73 (0.42, 1.24) | 0.243 | 14/365 | 1.36 (0.58, 3.17) | 0.480 |
| Race/ethnicity |  |  |  |  |  |  |
| Non-Hispanic Whites | 11/201 | Reference |  | 9/201 | Ref |  |
| Non-Hispanic Blacks | 29/176 | **3.32 (1.56, 7.07)** | **0.002** | 14/176 | **3.44 (1.32, 9.00)** | **0.012** |
| Hispanics | 25/224 | **3.44 (1.42, 8.34)** | **0.006** | 1/224 | 0.63 (0.07, 5.82) | 0.681 |
| Other | 0/88 | -<sup>a</sup> |  | 2/88 | -<sup>a</sup> |  |
| Ever smoker | 27/170 | **2.34 (1.23, 4.46)** | **0.010** | 10/170 | 0.82 (0.33, 2.01) | 0.659 |
| **Comorbidities** |  |  |  |  |  |  |
| Obesity | 19/128 | 1.47 (0.76, 2.83) | 0.250 | 5/128 | 1.09 (0.36, 3.33) | 0.881 |
| Diabetes | 34/170 | **3.31 (1.84, 5.96)** | **< 0.001** | 12/170 | 1.79 (0.75, 4.27) | 0.193 |
| Pure hypercholesterolemia | 7/20 | **3.77 (1.25, 11.36)** | **0.018** | 3/20 | -<sup>a</sup> |  |
| Asthma | 20/70 | **4.33 (2.18, 8.58)** | **< 0.001** | 3/70 | -<sup>a</sup> |  |
| COPD | 18/61 | **4.26 (1.87, 9.77)** | **0.001** | 5/61 | 1.29 (0.40, 4.13) | 0.667 |
| Chronic kidney disease | 22/81 | **5.63 (2.72, 11.64)** | **< 0.001** | 13/81 | **4.48 (1.81, 11.08)** | **0.001** |
| Cardiovascular disease | 50/341 | **5.59 (2.57, 12.14)** | **< 0.001** | 0/341 | -<sup>a</sup> |  |
| Neoplasm or history of neoplasm | 21/139 | 1.72 (0.89, 3.31) | 0.105 | 9/136 | 1.06 (0.42, 2.66) | 0.900 |
| Osteoarthritis | 19/105 | 2.01 (0.98, 4.11) | 0.057 | 8/105 | 1.10 (0.42, 2.89) | 0.854 |
| Vitamin D deficiency | 18/89 | **2.55 (1.28, 5.08)** | **0.008** | 5/89 | 1.08 (0.37, 3.17) | 0.889 |
| **Hematologic disorders** |  |  |  |  |  |  |
| Anemia | 39/176 | **5.19 (2.80, 9.65)** | **< 0.001** | 14/176 | **2.58 (1.05, 6.38)** | **0.040** |
| Coagulation defect | 17/55 | **4.67 (2.28, 9.57)** | **< 0.001** | 9/55 | **8.81 (3.11, 24.98)** | **< 0.001** |
| Thrombocytopenia | 11/37 | **5.14 (2.18, 12.09)** | **< 0.001** | 8/37 | **14.12 (4.54, 43.84)** | **< 0.001** |

Abbreviations: ICU, intensive care unit; OR, odds ratio; CI, confidence interval; COPD, chronic obstructive pulmonary disease; Ref, reference; n/N, cases/cases and non-cases in subgroup. Odds ratios calculated using multinomial logistic regression. Models adjusted for age, gender, race/ethnicity, and smoking.
-<sup>a</sup> odds ratios not computed.

### Factors Associated with Length of Hospital Stay

In adjusted analysis, male sex (β: 0.39, 95% CI: 0.16, 0.62) had longer length of hospital stay while Hispanics (β: -0.40, 95% CI: -0.74, -0.06) and participants of ‘Other’ race/ethnicity (β: -0.65, 95% CI: -1.16, -0.14) had shorted length of hospital stay than non-Hispanic Whites. Comorbidities such as diabetes (β: 0.50, 95% CI: 0.26, 0.74), asthma (β: 0.50, 95% CI: 0.20, 0.81), COPD (β: 0.45, 95% CI: 0.11, 0.79), cardiovascular disease (β: 0.40, 95% CI: 0.10, 0.70), and vitamin D deficiency (β: 0.47, 95% CI: 0.20, 0.75) were associated with longer length of hospital stay. Likewise, the hematological disorders (anemia [β: 0.38, 95% CI: 0.14, 0.62], coagulation defect [β: 0.57, 95% CI: 0.26, 0.88], and thrombocytopenia [β: 0.67, 95% CI: 0.30, 1.30]) were all associated with prolonged duration of hospital stay. (Table 5).

**Table 5:**
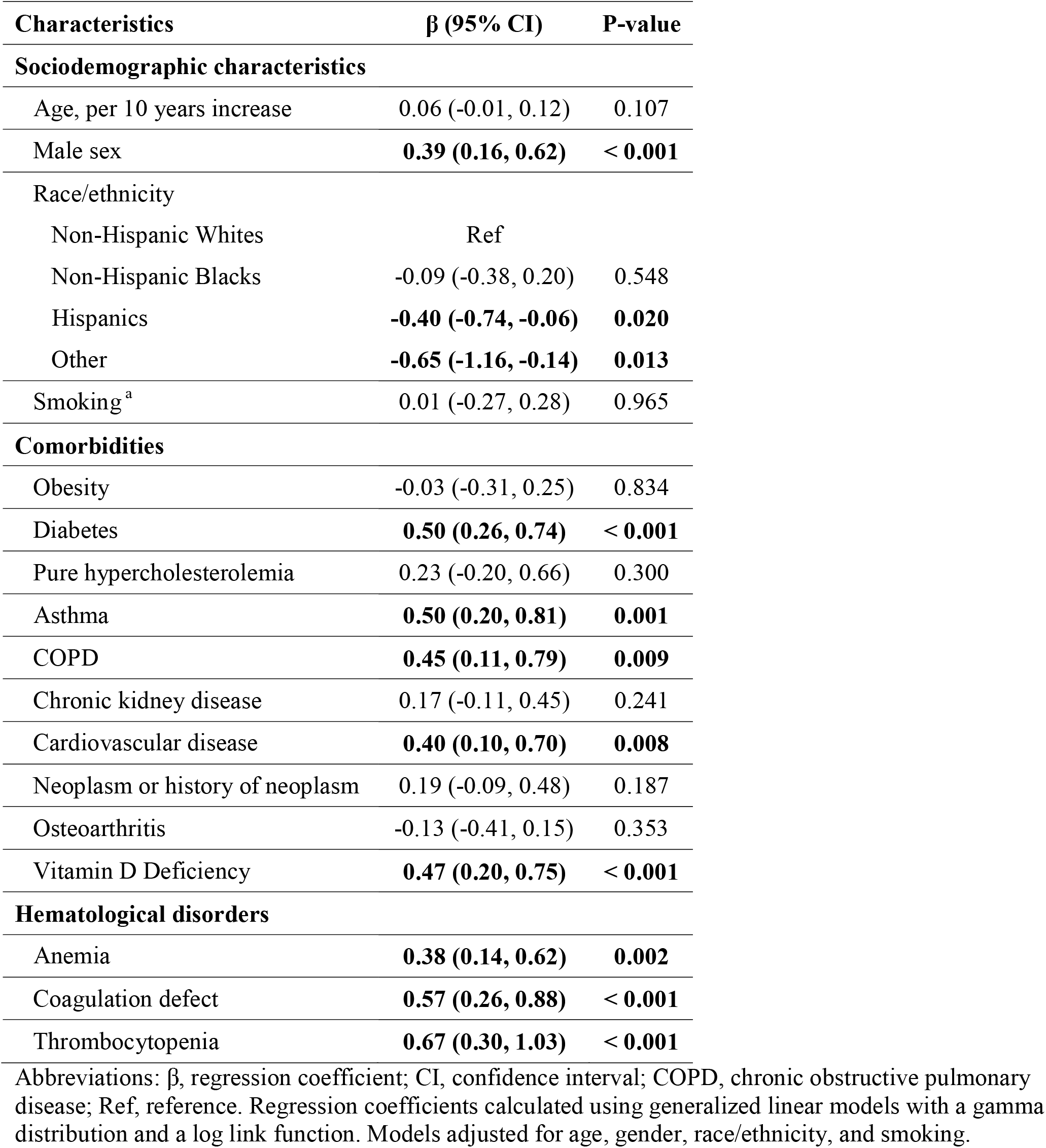
Factors associated with length of hospital stay in days (N = 191)

| Characteristics | $\beta$ (95% CI) | P-value |
| --- | --- | --- |
| **Sociodemographic characteristics** |  |  |
| Age, per 10 years increase | 0.06 (-0.01, 0.12) | 0.107 |
| Male sex | **0.39 (0.16, 0.62)** | **< 0.001** |
| Race/ethnicity |  |  |
| Non-Hispanic Whites | Ref |  |
| Non-Hispanic Blacks | -0.09 (-0.38, 0.20) | 0.548 |
| Hispanics | **-0.40 (-0.74, -0.06)** | **0.020** |
| Other | **-0.65 (-1.16, -0.14)** | **0.013** |
| Smoking<sup>a</sup> | 0.01 (-0.27, 0.28) | 0.965 |
| **Comorbidities** |  |  |
| Obesity | -0.03 (-0.31, 0.25) | 0.834 |
| Diabetes | **0.50 (0.26, 0.74)** | **< 0.001** |
| Pure hypercholesterolemia | 0.23 (-0.20, 0.66) | 0.300 |
| Asthma | **0.50 (0.20, 0.81)** | **0.001** |
| COPD | **0.45 (0.11, 0.79)** | **0.009** |
| Chronic kidney disease | 0.17 (-0.11, 0.45) | 0.241 |
| Cardiovascular disease | **0.40 (0.10, 0.70)** | **0.008** |
| Neoplasm or history of neoplasm | 0.19 (-0.09, 0.48) | 0.187 |
| Osteoarthritis | -0.13 (-0.41, 0.15) | 0.353 |
| Vitamin D Deficiency | **0.47 (0.20, 0.75)** | **< 0.001** |
| **Hematological disorders** |  |  |
| Anemia | **0.38 (0.14, 0.62)** | **0.002** |
| Coagulation defect | **0.57 (0.26, 0.88)** | **< 0.001** |
| Thrombocytopenia | **0.67 (0.30, 1.03)** | **< 0.001** |

Abbreviations: $\beta$ , regression coefficient; CI, confidence interval; COPD, chronic obstructive pulmonary disease; Ref, reference. Regression coefficients calculated using generalized linear models with a gamma distribution and a log link function. Models adjusted for age, gender, race/ethnicity, and smoking.

## DISCUSSION

In this racially and ethnically diverse study population of COVID-19 patients, the sociodemographic characteristics associated with hospitalization and/or disease severity included older age, non-Hispanic Black or Hispanic race/ethnicity (compared to non-Hispanic Whites). Among the studied comorbidities, diabetes, pure hypercholesterolemia, asthma, COPD, chronic kidney disease, cardiovascular disease, osteoarthritis, and vitamin D deficiency were risk factors associated with hospitalization and/or severe COVID-19. Hematological disorders such as anemia, coagulation defects, and thrombocytopenia were associated with increased odds of both hospitalization and disease severity.

This report is one of a few epidemiological studies investigating the risk factors for disease severity in a racially and ethnically diverse sample of COVID-19 patients. In the UK, three studies analyzed data from the national Biobank and, consistent with our findings on non-Hispanic Blacks, found excess hospitalizations in Blacks and Asians compared to Whites.^23-25^ In the US, five studies conducted among hospitalized COVID-19 patients observed an overrepresentation of non-Hispanic Blacks, with prevalences ranging from 51.0% to 83.2%.^9,12,25-27^ However, the studies found that non-Hispanic Blacks were not at higher risk of severe disease or death compared to non-Hispanic Whites among hospitalized COVID-19 patients.^9,12,26-28^ A large U.S. Department of Veterans Affairs study with 5,630 COVID-19 patients reported that non-Hispanic Blacks and Hispanics were more likely to test positive for COVID-19 than non-Hispanic Whites, but were not at higher risk of 30-day mortality.^20^ Of all of these studies, only one examined length of hospital stay as an outcome and it found no difference by race or ethnicity.^12^ Our results also suggested that although non-Hispanic Blacks and Hispanics were more likely to be hospitalized and to have severe COVID-19 than non-Hispanic Whites, length of hospital stay was not different between non-Hispanic Whites and non-Hispanic Blacks. It was even shorter in Hispanics and patients of ‘Other’ race/ethnicity than in non-Hispanic Whites. It is possible that the higher death rates in non-Hispanic Blacks could have affected the length of hospital stay and explained the non-significant difference with non-Hispanic whites. It is unclear, however why hospital stay was shorter in Hispanics and patients of ‘Other’ race/ethnicity.

Our results confirmed previous reports that older age and smoking as well as comorbidities such as diabetes, cardiovascular diseases, chronic kidney, COPD, coagulation defect, and thrombocytopenia are associated with hospitalization and/or severe COVID-19.^6,9,27,29,30^ Consistent with the longer length of hospital stay noted in men in our analysis, it has been suggested that male sex may predispose to more severe COVID-19 due to hormonal factors.^11,26^ Males are known to have higher activity of angiotensin-converting enzymes 2 (ACE2), the receptors of which serve as an entry point for the SARS-CoV-2 in the alveolar epithelial cells and this could explain the increased risk of infection and disease severity in men previously reported in the literature.^31^ Another hypothesis attributes the sex difference in COVID-19 severity to the transmembrane protease serine 2 (TMPRSS2) gene that is influenced by androgen/estrogen stimulation and facilitates the fusion of the viral and cellular membranes.^32^

Although we found no association between obesity and severe COVID-19, other studies observed that morbid obesity (body mass index ≥40) was a risk factor of disease severity.^11,27^ With regards to patients with neoplasm or a history of neoplasm, they were not found to have higher COVID-19 severity in our study. One Chinese study performed on 1,590 COVID-19 cases observed that the 18 patients with a history of cancer had a higher risk of disease severity in their crude analysis.^32^ However, these patients were older (63.1 years versus 48.7 years) and more likely to have been smokers (22% versus 7%) than the COVID-19 patients without a cancer history.^33^ A recent meta-analysis summarizing the existing literature on cancer and COVID-19 severity concluded that the current evidence on this association remains inconclusive.^34^ Likewise, the evidence of an association between asthma and COVID-19 severity is scarce and the previous studies were underpowered as they included few asthma cases among COVID-19 patients.^6,15^ Consistent with our findings, a recent analysis of UK biobank data concluded that asthma was associated with severe COVID-19 among 641 infected patients and that this association was driven by non-allergic asthma.^17^

Our epidemiological study is also the first to investigate the association of conditions such as vitamin D deficiency, pure hypercholesterolemia, osteoarthritis, and anemia with severe COVID-19 An analysis of UK biobank data examined the serum levels of vitamin D and the risk of infection with COVID-19 and found no association, while another study from the University of Chicago showed an increased risk of COVID-19 infection associated with vitamin D deficiency.^18,35^ However, none of the studies investigated the association of vitamin D with disease severity among patients who were already infected with SARS-CoV-2.^18,35^ In an ecological study, mortality rates among COVID-19 patients were compared among developed nations and it was suggested that countries with higher prevalence of vitamin D deficiency had higher mortality rates.^36^ Yet, ecological studies do not include patient-level data and suffer from ecological fallacy, which is the assumption that factors associated with the national disease rates are associated with disease in individual patients.^37^ Nevertheless, there is reason to investigate Vitamin D status as a factor in disease progression. Vitamin D is well-known for its immunoregulatory properties; it can increase cellular immunity by stimulating antimicrobial peptides and could oppose cytokine storms induced by the innate immune responses.^38,39^ No published epidemiological study has also investigated whether high cholesterol is associated with higher risk of COVID-19 severity, although hypercholesterolemia is a major contributor to atherosclerosis and cardiovascular disease, which itself, is associated with severe COVID-19.^40^ Furthermore, high cholesterol might enhance the replication of SARS-CoV-2 in endothelial cells potentially causing acute vascular injury and triggering coagulopathies.^41^ No study has also examined the relationship of osteoarthritis and anemia with severe COVID-19. Osteoarthritis is a common joint disease, particularly prevalent in older adults; it is associated with various comorbidities and with an increased risk of mortality from cardiovascular diseases, diabetes and renal diseases.^42,43^ Anemia affects 5.6% of all Americans, but this prevalence is increased to more than 10% in Americans aged 65 or older.^44,45^ It is well-known to be a factor for poor prognosis in respiratory diseases such as COPD.^46^ In our study however, it is unclear whether anemia is a factor for severe COVID-19 or a consequence of the inflammation and cytokine production caused by the infection and leading to hemolytic anemia.^46^

Our study has limitations. It included only patients from a single health system in the Midwest of the U.S. and may not be generalizable to the overall American population. Our study design was observational; therefore, temporality and causality between certain factors (anemia, coagulation defects, and thrombocytopenia for instance) and COVID-19 severity cannot be established. Smoking status was missing for 22.8% of the COVID-19 participants. Vitamin D deficiency was defined using electronic medical records and data on the serum vitamin D levels were not available. Nonetheless, our study has major strengths, it was conducted in a large and racially/ethnically diverse sample of COVID-19 patients. It also included factors formerly understudied in COVID-19 such as vitamin D deficiency, pure hypercholesterolemia, osteoarthritis, and anemia.

In conclusion, the present study confirms previous reports that older age, non-Hispanic Black or Hispanic race/ethnicity, smoking, diabetes, COPD, chronic kidney disease, cardiovascular disease, coagulation defects, and thrombocytopenia are associated with severe COVID-19. It also identifies severe COVID-19 factors not previously reported such as vitamin D deficiency, pure hypercholesterolemia, osteoarthritis, and anemia. Our results are of public health relevance and have implications in the prioritization of COVID-19 patients at risk of severe disease and in the implementation of interventions to mitigate negative outcomes. Future studies should also evaluate whether vitamin D supplementation in COVID-19 patients with vitamin D deficiency might improve disease prognosis.

## Data Availability

The datasets and SAS codes used for data analysis are available only for the purpose of reproducing the study findings upon reasonable request.

